# Remote recruitment for Essential Coaching for Every Mother during COVID-19

**DOI:** 10.1101/2021.03.09.21253071

**Authors:** Justine Dol, Gail Tomblin Murphy, Douglas McMillan, Megan Aston, Marsha Campbell-Yeo

## Abstract

**Background:** With a decrease in in-person support and increase in perinatal mental health concerns during the coronavirus pandemic, innovative strategies, such as mHealth, are more important than ever. However, due to physical distancing recommendations, recruitments for perinatal research needs to shift. The objective of this study is to desire the process evaluation of recruitment and retention of women for an mHealth pre-post intervention study for *Essential Coaching for Every Mother*.

**Methods:** Three methods were used for recruitment: social media, posters in hospital, and media outreach. First time mothers were eligible for enrollment antenatally (37+ weeks) and postnatally (<3 weeks). Eligibility screening occurred remotely via text message. Outcomes were days to recruit 75 participants, eligibility vs. ineligibility rates, dropout and exclusion reasons, survey completion rates, perinatal timing of enrollment, and recruitment sources.

**Results:** Recruitment ran July 15^th^-September 19^th^ (67 days) with 200 screened and 88 enrolled, 70% antenatally. It took 50 days to enroll 75 participants. Mothers recruited antenatally (n=53) were more likely to receive all intervention message (68% vs. 19%). Mothers recruited postnatally (n=35) missed more messages on average (13.8 vs. 6.4). Participants heard about the study through family/friends (31%), news (20%), Facebook groups (16%), Facebook ads (14%), posters (12%), or other ways (7%).

**Conclusion:** Antenatal recruitment resulted in participants enrolling earlier and receiving more of the study messages. Word of mouth and media outreach were successful, followed by advertisement on Facebook. Remote recruitment was a feasible way to recruit for *Essential Coaching for Every Mother*.

---

Irrespective of a pandemic, mothers living in Nova Scotia and beyond face gaps in access to information and often struggle to find adequate support during the postpartum period, defined as the first six weeks after birth.^1–3^ These gaps may be magnified during the coronavirus pandemic and may significantly impact the transition for new mothers.^4,5^ Compliance with physical distancing recommendations contribute to mothers isolating at home, being physically isolated from not only health providers, but also from their extended family and support systems.^4^ In Nova Scotia, all public health drop-ins were closed indefinitely, there was a reduction in in-person healthcare support, and midwifery-led home births and home visits were temporarily deferred during the coronavirus peak from March to May 2020.^6^ This significantly differed from pre-coronavirus procedures, where mothers were recommended to have a postnatal contact shortly after birth by a public health nurse^7^ and mothers frequently engaged in visits with family, friends, or new parent groups.^8,9^ Emerging evidence shows that the pandemic has resulted in 37-54% of mothers experiencing perinatal depression and 57-72% experiencing symptoms of perinatal anxiety.^10,11^ With the sudden decrease in in-person support and the increase in perinatal mental health concerns, innovative strategies, such as mHealth, are more important than ever to offer information and support during the postpartum period.

Prior to the coronavirus outbreak, the *Essential Coaching for Every Mother* program was developed to send daily text messages to mothers during the immediate six-week postpartum period.^12^ However, given the requirement of physical distancing and limitations on the number of visitors in hospital, exploration was needed on the ability to recruitment remotely rather than the traditional, in-person approach for a planned randomized control trial. Therefore, this study sought to explore the feasibility of remote recruitment of pregnant and postpartum women for the *Essential Coaching for Every Mother* pre-post intervention study.

## Methods

### Research Design

This study uses a cross-sectional pre-post design.

### Study Population & Sample Size

Between 2017 and 2019 at the IWK Health Centre, 4,055 primiparous women gave birth.^13^ Targeting recruitment over three months, a potential population of approximately 500 mothers would be available. To determine feasibility of recruitment, the goal was to recruit at least 15% of this sample (n=75) within three months. The recruitment goal for the randomized controlled trial is 140 participants,^14^ so this estimated sample size was approximately half of what would be required.

To participate, women must (1) have given birth to their first baby at IWK Health and live in Nova Scotia; (2) have daily access to a mobile phone with text message capabilities; (3) be over 18 years of age; and (4) speak and read English. Women were eligible to enrol antenatally if they were at least 37 weeks pregnant and had not yet given birth. The antenatal time limitation was set to ensure participants would deliver within the three-month recruitment period. Women were eligible to enrol postnatally up to 21 days following the birth of their child. The postpartum limit was set to ensure there was a least a three-week gap between baseline and 6-week follow-up surveys and to ensure participants received enough of the message to provide evaluative feedback.

### Intervention

*Essential Coaching for Every Mother* is a six-week postpartum text message program that was previously developed in consultation with postpartum mothers and healthcare providers with the goal of improving women’s psychosocial outcomes.^12^ With the outbreak of COVID-19 in early 2020 and the readiness of the *Essential Coaching for Every Mother* program to fill the sudden gap in postpartum support, a decision was made to modify the program to be offered immediately. Given that *Essential Coaching for Every Mother* was developed prior to the coronavirus outbreak but not previously implemented, some modifications were necessary. To ensure that the revised content of *Essential Coaching for Every Mother* was appropriate and acceptable, the modified messages were piloted with mothers and postpartum healthcare providers who participated in the original development of the program. Messages were updated using the Government of Canada and World Health Organization guidelines around mother-infant care and coronavirus^15,16^ and followed the Government of Nova Scotia public health guidelines during the coronavirus pandemic.^17^

Nine messages were modified from the original program to include information related to the coronavirus, of which four messages were collapsed into two, and five messages were added to the program. This resulted in a total of 56 messages, with messages provided two or three times per day during the two weeks, and daily for the remaining four weeks. Figure 1 provides an example of two messages included. The first message of *Essential Coaching for Every Mother* is designed to start the evening of the second day after giving birth. If a participant signed up beyond this time frame, they started the messages based on when they delivered.

**Figure 1.**
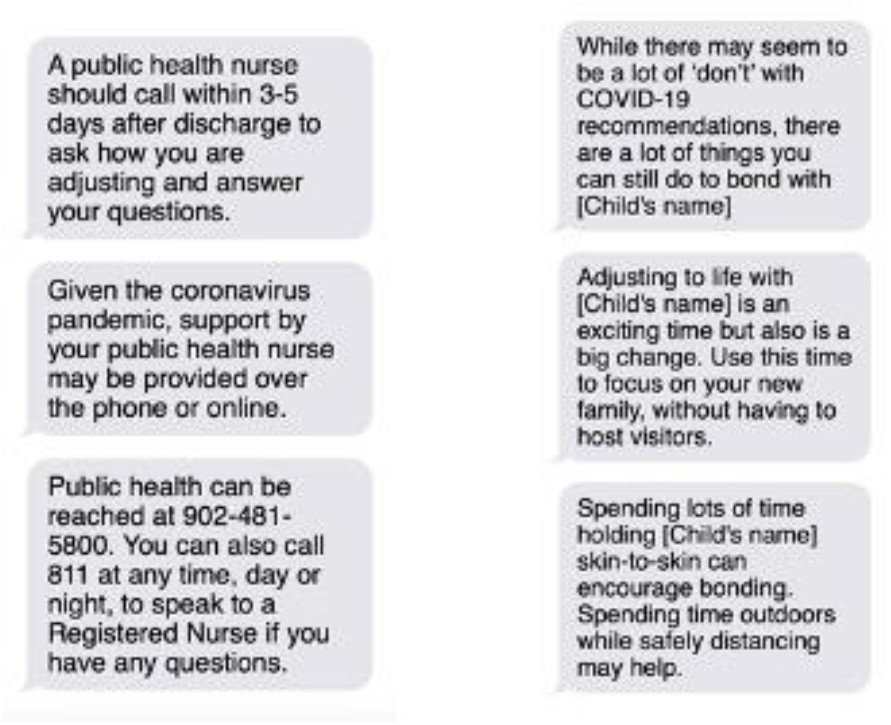
Examples of Essential Coaching for Every Mother messages relevant to COVID-19. The first is a message that was revised to reflect the current standard of care in postnatal follow-up by a public health nurse which increased from within 48 hours to 3-5 days and may occur over the phone or online rather than in person. The second message is an example of a message that was added related specifically to postnatal care during COVID-19.

### Recruitment Procedures

Three primary methods of recruitment were used: social media, posters in the hospital, and media outreach. First, social media advertisements were used including paid Facebook and Instagram advertisements, sharing in relevant Facebook groups, and tweets on Twitter. Social media outreach and paid advertisements started on July 15^th^ and ran until August 16^th^, 2020. Second, posters were placed in the IWK Health Perinatal Clinic and in each room on the Family Newborn Unit. Posters were placed on August 5^th^ and taken down on September 15^th^. Finally, media interviews also occurred with the first author after a media release was published by IWK Health on August 5^th^, 2020.

All eligibility screening occurred remotely via text message through the TextIt platform^18^ with interested participants initiating contact. Pregnant women started the recruitment process by texting ‘pregnant’ to the study number and proceeded through the antenatal eligibility screening process. Eligible mothers were instructed to text ‘delivered’ after giving birth to be enrolled in the study. Women received reminder messages to text ‘delivered’ at 39 weeks, 40 weeks, 41 weeks and 42 weeks if they had not yet enrolled or withdrawn. Mothers who were deemed ineligible as part of the antenatal screening due to being less than 37 weeks were sent a message to remind them to text ‘pregnant’ if they were still interested. This occurred until August 27^th^ when the number of interested and enrolled participants was beyond the desired 75 participants.

Postpartum women who initiated contact using ‘birth’ proceeded through the postpartum eligibility flow. Once deemed eligible, postpartum participants and antenatal women who texted ‘delivered’ completed the same flow to be enrolled in the study and start receiving messages based on their delivery date. During this phase, details about newborn’s name, preferred gender pronoun, date of birth, mother’s name, and preference for breastfeeding or formula messages were collected. This was used to personalize the messages with names and ensure messages were sent based on child’s age and preference for breastfeeding or formula messages.

Participants were asked to complete a consent form and survey at baseline (survey #1) and once the messages ended at 6-weeks (survey #2). Participants were reminded about the surveys via text message six times (every 2 days) at each timepoint or until they completed the full survey. After 14 days, participants who had yet to complete the full survey were sent an email or text as a final reminder and it was assumed to be incomplete if a participant did not complete after this point. Participants who completed the full survey received a $20 electronic gift card at each survey timepoint. Figure 2 outlines of study enrollment and participation flow.

**Figure 2.**
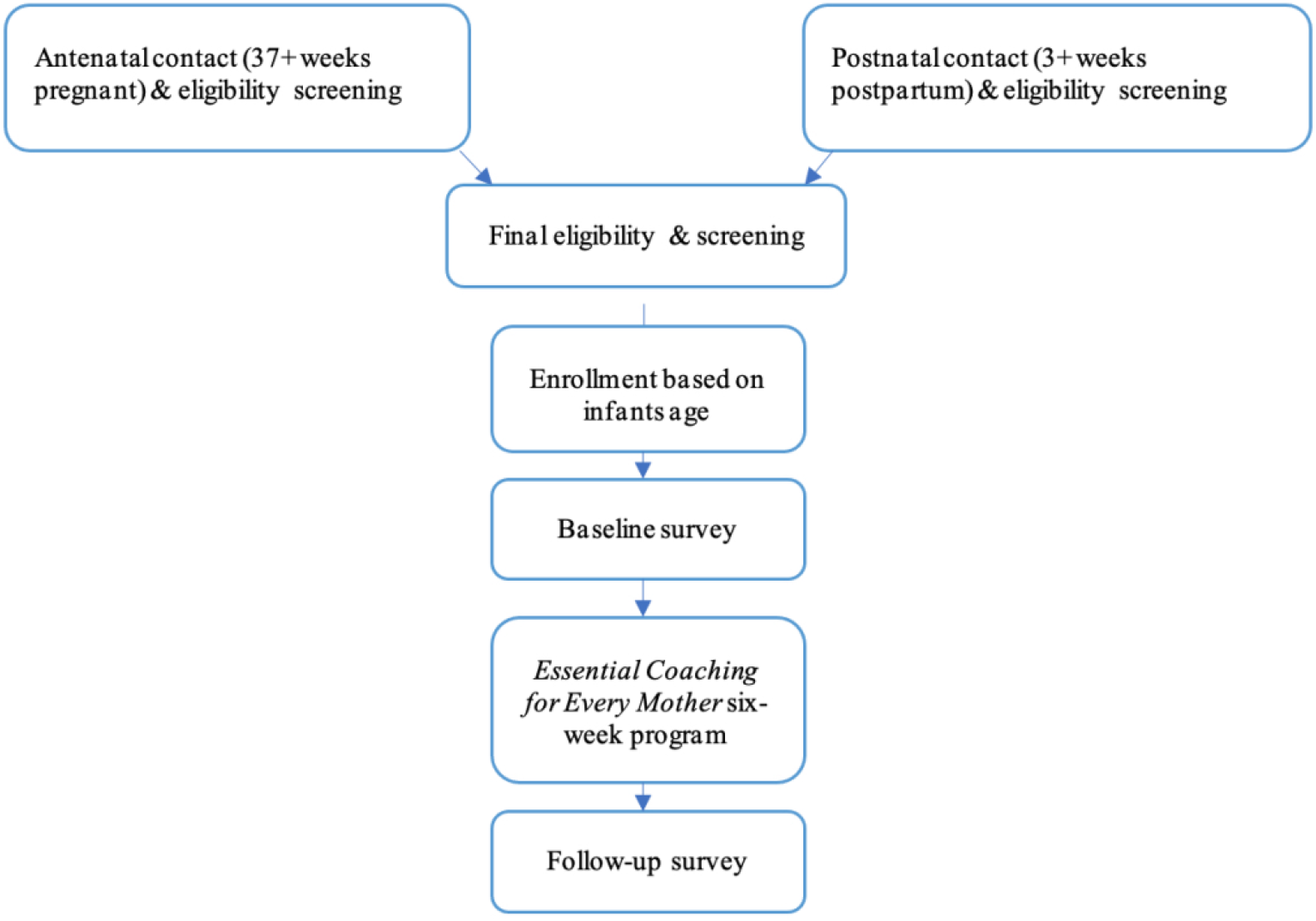
Study enrollment and participation flow diagram

While messages were only one-way, if a mother responded with a question of clarification during recruitment or related to the survey, the first author responded to provide clarification only. No responses to the *Essential Coaching for Every Mother* messages occurred.

### Intervention Procedures

TextIt^18^ was used as the platform to develop the message flows and capture participant contact information. It was used in conjunction with Twilio^19^ the server that sent and received the messages. Survey data were collected via REDCap.^20^ Contact information was collected via TextIt which was kept separate from survey data collected via REDCap. Ethics approval was obtained by the IWK Health Centre (#1024984). All women provided written, online consent.

To determine feasibility, data on implementation extent was collected via output data available through the TextIt platform as well as REDCap. Specifically, the following outcomes were of interest: (1) number of days required to recruit at least 75 participants; (2) eligibility vs. ineligibility rates and reasons; (3) dropout, exclusion, and baseline survey completion rates; (4) enrollment rates based on antenatal or postnatal recruitment; and (5) recruitment sources. For the outcomes identified above, the following information was used: Days required to recruit participants was the number of days from start of study to enrollment of 75 participants, total number of participants enrolled, and time required for recruitment. Eligibility vs. ineligibility rates was the number of individuals who were eligible and enrolled in the study vs. the number of individuals who contacted but were not eligible (comparing both antenatal and postpartum ineligibility and reasons). Dropout & survey completion rates was the number of participants who withdrew or were excluded from the study and when (% completion, timing of withdraw). Enrollment based on antenatal vs. postnatal recruitment compared number and timing of enrollment (number of messages received, infants age at enrollment, time of day of initial contact). Recruitment sources were participant’s self-reported source of where they heard about the study.

### Analysis

Descriptive and summative analysis of TextIt and REDCap event data was used to examine the frequency and proportion of outcomes above.

## Results

### Participants

Of the 80 women who completed the demographic survey, they were on average 30.8 years of age (standard deviation [SD]=4.7). The sample was quite homogeneous – 98.8% had singleton births, 91.3% identified as heterosexual, 87.5% were white, and 93.8% were either married or common-law. Over half (51.8%) had a household income over $100,000CAN. Women were on average 39.3 weeks pregnant when they gave birth (SD=1.5 weeks, range=33.5-42 weeks).

### Timing required to recruit participants

The study opened on July 15, 2020, and closed September 19, 2020. Over this 67 days, 96 participants were enrolled in the program and were assigned a study identification number. Timing to enroll 75 participants (our initial target) took 50 days.

### Eligibility vs. ineligibility

A total of 200 ‘pregnant’ or ‘birth’ messages were sent to the study contact number by potential participants during the recruitment period. Figure 3 outlines the perinatal timing and reasons for ineligibility. 140 participants initiated contact antenatally and 60 initiated contact in the postpartum period. For the antenatal participants, 30 were not eligible, seven were not interested, and 45 were excluded not being based in Nova Scotia. For the postpartum period, 20 were not eligible and two were not interested.

**Figure 3.**
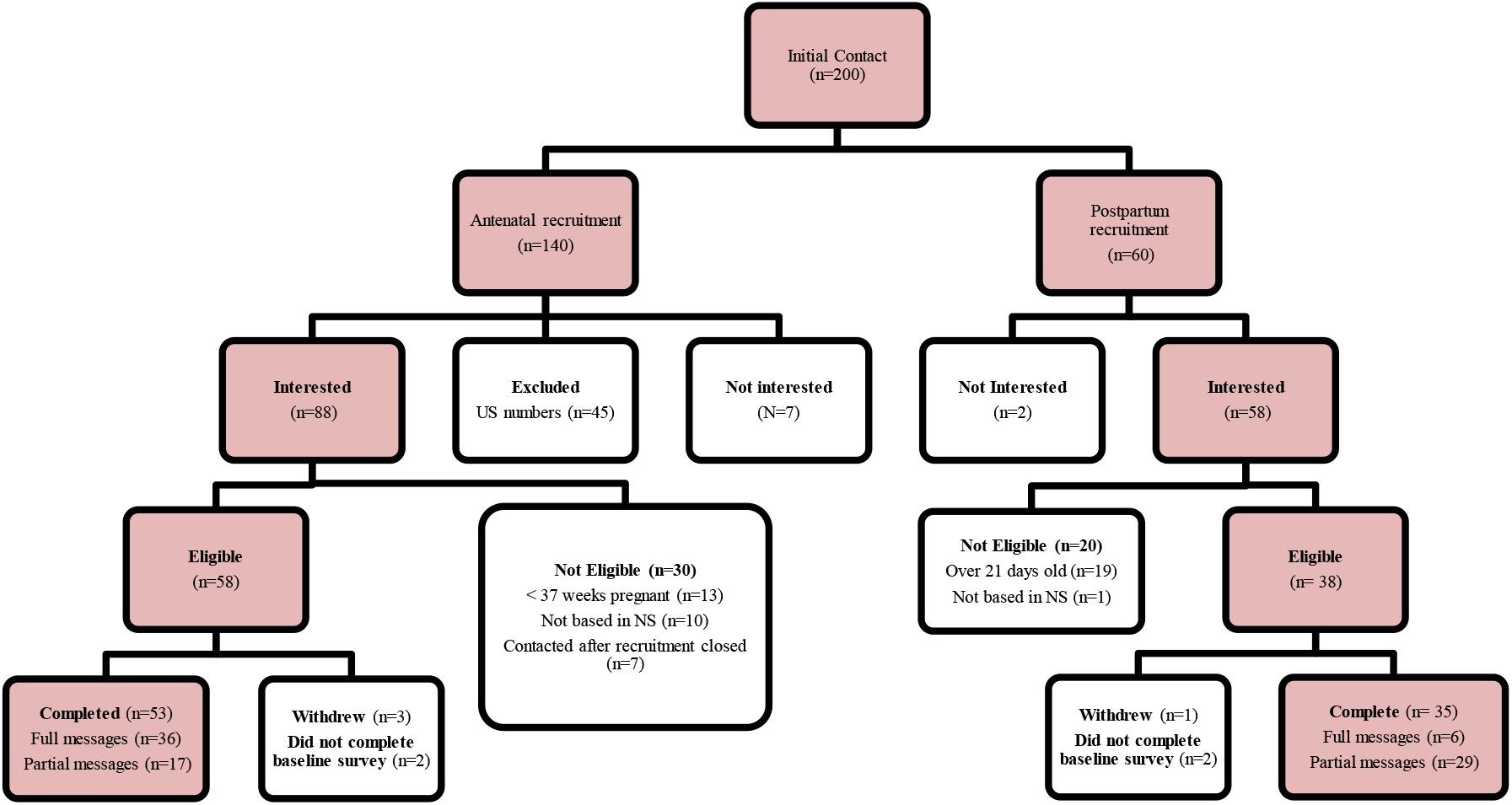
Enrollment flow

We excluded 45 contacts who were not based in Nova Scotia and were using a United States (US) number. We initially thought this could be individuals who are temporarily residing in Canada since they provided valid Nova Scotia postal codes in the demographic questionnaire. However, upon further analysis, we believe these were not actual mothers from the US wanting to participate due to a discrepancy in the standardized questionnaire responses with extremely high scores on these measures (well beyond the standard mean) and quick completion time of their REDCap surveys (immediately after they enrolled and on their delivery date), suggesting these were not actual participants. When analyzing location through Twilio, the US had a predominant send/receive rate, providing further evidence that these respondents were not in Canada. Therefore, with the triangulation of these findings, it was deemed that these responses are not actual potential participants and if they were, they were not residing in Nova Scotia as required by the study protocol, and thus were excluded.

### Dropout & survey completion rates

Of the 96 enrolled participants, four withdrew from the program after receiving 0, 5, 6 and 9 messages respectively (Mean [M]=4, SD=3.7). Three of the participants who withdrew enrolled in the postpartum period and one enrolled antenatally. As none of the participants who withdrew completed the baseline survey, we were unable to determine if these participants were different from those who completed the program. Four participants did not complete any aspect of the baseline survey, thus were excluded from the analysis.

Therefore, the study had a total of 88 participants who did not opt out and who completed at least some of the baseline survey. Ninety percent (90.1%) of participants completed the full baseline survey, on average 5.0 days from enrollment (Median = 3 days, SD=5.3 days, range 0-19 days). Nine percent (n=8) did not complete baseline survey in full – on average, participants completed 56.25% of the survey (range: 25%-75%).

### Timing of recruitment

Of the 88 participants who were enrolled, 42 (47.7%) received full messages. Of these, 36 were antenatally recruited and six were recruited postnatally. Late enrollment during the antenatal period resulted in missing on average 6.4 messages (SD=6.2) whereas late enrollment during the postpartum period resulted in missing on average 13.8 messages (SD=10.6).

### Recruitment sources

Among the 80 participants who completed the full survey, 30.5% (n=25) heard about the study through friends or family and 18.3% (n=15) heard about it on the news. Recruitment via Facebook was also successful, with a quarter of participants reached through the social media platform – 14.6% via Facebook groups, 13.4% via Facebook advertisements, and 1.2% via Facebook Marketplace. No participant reported hearing about it through Instagram or Twitter. Posters in the hospital was the source of recruitment for 14.6% of participants, with 7.6% saying other (including doula, social media broadly, hospital website, and no response).

For paid Facebook advertisements, a total of $215.77 Canadian was spent, which equals a cost of $19.62 per enrolled participants who indicated this a primary recruitment method. However, this may not be accurate as this does not consider whether any friends or family heard about the study through paid advertisements.

## Discussion

This study describes the remote recruitment of *Essential Coaching for Every Mother* as a pilot pre-post intervention study. The online and remote recruitment of pregnant and postpartum women for a pre-post intervention study for *Essential Coaching for Every Mother* was a success as we were able to recruit over our target of 75 participants within 50 days, with recruitment suspended within 67 days due to significant interest. This suggests that mothers were interested in receiving information during the postpartum period, which may have been enhanced due to the pandemic.

Emerging evidence shows that the pandemic has resulted in 37-54% of mothers experiencing perinatal depression and 57-72% experiencing symptoms of perinatal anxiety,^10,11^ suggesting that a preventative mHealth program for mothers could have a positive effect on mothers postpartum adjustment and experience. Given the growing evidence of the mental health consequences of physical distancing recommendations,^21^ particularly during an intensely vulnerable period as is the postpartum period, having evidence-based information provided via text message may help cover this gap. Digital health during COVID-19 has the potential to bridge the healthcare service gap while maintaining physical distancing recommendations.^22,23^

We found that mothers who were recruited antenatally received more of the study messages than participants who were recruited postnatally, with the latter missing on average 7.4 messages more. As participants who initiated contact during the antenatal period were sent reminder messages starting at 39 weeks, they were more likely to enroll earlier than mothers who had already delivered. No mother who expressed interest during the antenatal period and was deemed eligible failed to enroll. Thus, antenatal recruitment may be a more efficient way to target recruitment for the larger clinical trial to ensure mothers receive as much of the program as possible. Additionally, given the delay in baseline survey completion after delivery, shifting to have participants complete the baseline survey upon enrollment and prior to delivery may result in more timely completion than during the postpartum period.

Looking at the direct recruitment methods, the most successful approach was promotion through Facebook. Both advertisement through mother-focused Facebook groups and paid advertisements were similarly effective. This finding is supported by previous systematic reviews which found that Facebook recruitment was an effective way to reach participants for health research.^24,25^ Within our study, we also found that sharing the study in the media and news reached 17% of participants and posters in the hospital reached 13.6% of participants, which suggests that using a multi-pronged approach to recruitment is more efficient than solely using social media.

## Limitations

Despite the successes, there were some challenges in recruitment. First, most participants heard about the study through family and friends, but it is unclear how these family and friends heard about it. Additionally, we were unable to gather how mothers who contacted us but did not enroll in the study heard about it. Both of these factors limit the interpretation of recruitment source analysis. A second challenge was the high potential for people to misuse the self-identification of eligibility screening which occurred exclusively via text message. This occurred in relation to the large number of US-based numbers. We hypothesize that someone(s) had been completing the eligibility screening and baseline questionnaire to gain access to the honorarium. While TextIt cannot limit to provincial locations, we continued to monitor recruitment closely to ensure we identified any issues related to this through regularly monitoring of area codes. This may have potentially excluded individuals who were residing in Nova Scotia but had US numbers, this was required to ensure safety and adherence to study protocol inclusion criteria. In terms of the sample being primarily white and middle-upper income, this was recognized as a limitation and is consistent with online recruitment.^24^ Further work should have a more direct focus on collecting a diverse sample.

## Conclusion

Despite these challenges, this study found that *Essential Coaching for Every Mother* was able to successfully use remote recruitment of pregnant and postpartum women for a pre-post intervention study using a variety of recruitment sources. Findings from this study will be applied in the randomized controlled trial.^14^

## Data Availability

Data is not available.

